# Rural Diets Under Pressure: Food Environments and their Influence on Food Choice in South Asia

**DOI:** 10.1101/2025.01.04.25319997

**Authors:** Alka Chauhan, Samuel Scott, William Joe, Nanda Kumar Maharjan, Purnima Menon, TAFSSA collaborators, Suman Chakrabarti

**Author notes:** Correspondence to Suman Chakrabarti, Associate Research Fellow, International Food Policy Research Institute, Washington DC, USA.

## Abstract

The rapid South Asia rural transformation, driven by globalization and industrialization, has introduced a complex interaction between traditional and modern food systems. This study characterizes rural food environments in five districts across Bangladesh, India, and Nepal, focusing on how affordability, availability, accessibility, desirability, and convenience shape dietary choices and quality. Through extensive household and market surveys, we find rural diets characterized by low intake of healthy foods and moderate to high consumption of unhealthy options, influenced by a lack of affordability and the desirability and widespread availability of cheap ultra-processed products in rural markets. Snacking plays a significant role in shaping dietary patterns, promoting both dietary diversity and unhealthy food consumption. These findings provide essential insights for designing interventions tailored to rural food systems, supporting efforts to improve nutrition and health outcomes in rapidly changing markets.

## Introduction

Globalization and industrialization are reshaping food systems, particularly in low- and middle-income countries (LMICs), through a complex transformation of food environments ^1,2^. Food environments encompass “the interface where people interact with the wider food system to acquire and consume foods” ^3^. This interface includes physical, economic, policy, and sociocultural dimensions that collectively shape dietary behaviors. The physical dimension involves food source availability and accessibility; economic aspects cover food prices and affordability; policy elements include regulations and interventions; while sociocultural dimensions encompass cultural eating norms and marketing influences ^2,4^. Understanding these interconnected dimensions is crucial for addressing nutrition challenges and developing effective interventions to promote healthy eating behaviors.

LMICs have larger rural populations and are experiencing rapid transformations driven by economic growth, market integration, and technological advancements. As industrial food production expands, packaged food manufacturers capitalize on traditional distribution networks, ultimately increasing the accessibility of processed foods in rural areas ^5,6^. Concurrently, the rise in modern retail and household income has catalyzed an expansion of food market landscapes. This expansion brings a mix of healthier food options alongside a surge in the availability of ultra-processed products ^7,8^.

South Asia, home to the world’s largest rural population and significant agricultural activity, is at the center of these transformations. South Asian rural food markets present a unique dichotomy where traditional food systems coexist with an increasingly prominent influx of processed food options ^9,10^. While urban markets have long been saturated with diverse choices, rural markets are rapidly closing this gap, with western fast food and industrialized products penetrating even the most remote regions ^8,9^. Experience from middle- and high-income countries suggests that local food environments in LMICs are likely to be shaped by the influence of global food industry giants that have long neglected consumer health. This transformation reveals a disconnect between food availability and dietary quality, as the increased availability of diverse food options, including ultra-processed and unhealthy foods, does not necessarily translate into better health outcomes ^11,12^.

Despite emerging rural markets, existing research has largely concentrated on urban settings and high-income countries, resulting in a substantial gap in understanding rural food systems in LMICs and designing effective interventions tailored to their needs. This gap encompasses several essential dimensions, including the need to address attributes that shape dietary outcomes, methodological challenges in capturing the complexities of local food environments, and the lack of accurate and timely data on diets and markets ^13^. For instance, in South Asia, local market structures such as *haats* (temporary markets) and *mandis* (organized markets) exemplify the diverse systems that influence food environments, yet these remain underexplored in the context of rural dietary behaviors. As LMICs undergo rapid urbanization, this transition presents a critical opportunity to study rural areas and implement interventions before they develop into urban centers characterized by unhealthy food environments.

This research addresses critical gaps by comprehensively examining rural food environments in selected districts across Bangladesh, India, and Nepal. The study seeks to answer two key questions: (1) What are the defining characteristics of local food environments in rural South Asia? and (2) How do attributes of the food environment—affordability, availability, accessibility, desirability, and convenience—influence food choices and diet quality?

## Methods

### Data

The Transforming Ari Food Systems in South Asia (TAFSSA) survey was conducted between February and May 2023 across five locations: Nalanda district in Bihar, India (N=1000 households); Surkhet (N=500) in Lumbini Province and Banke (N=500) in Karnali Province, Nepal; and Rangpur (N=1000) in Rangpur Division and Rajshahi (N=1000) in Rajshahi Division, Bangladesh. The household surveys are representative at the rural district level. Respondents included one adult female (aged 20+), one adult male (aged 20+), and one adolescent (aged 10–19) per household. To assess food environments, community market surveys were conducted in the same villages as the household surveys. In each village, six retailers and three restaurants were randomly selected and interviewed regarding vendor information, customer profiles, sales, essential goods, prices, supply chains, food waste, infrastructure, hygiene, and marketing practices. The study protocol, informed consent forms, and questionnaires were approved by the Institutional Review Boards (IRBs) of the International Food Policy Research Institute (Washington, DC, USA), the Institute of Health Economics (University of Dhaka, Bangladesh), Centre for Media Studies (Delhi, India), and the Nepal Health Research Council (Kathmandu, Nepal). Additional details are available in supplemental methods (S1), with indicator definitions and constructions provided in **Table 1**.

**Table 1.**
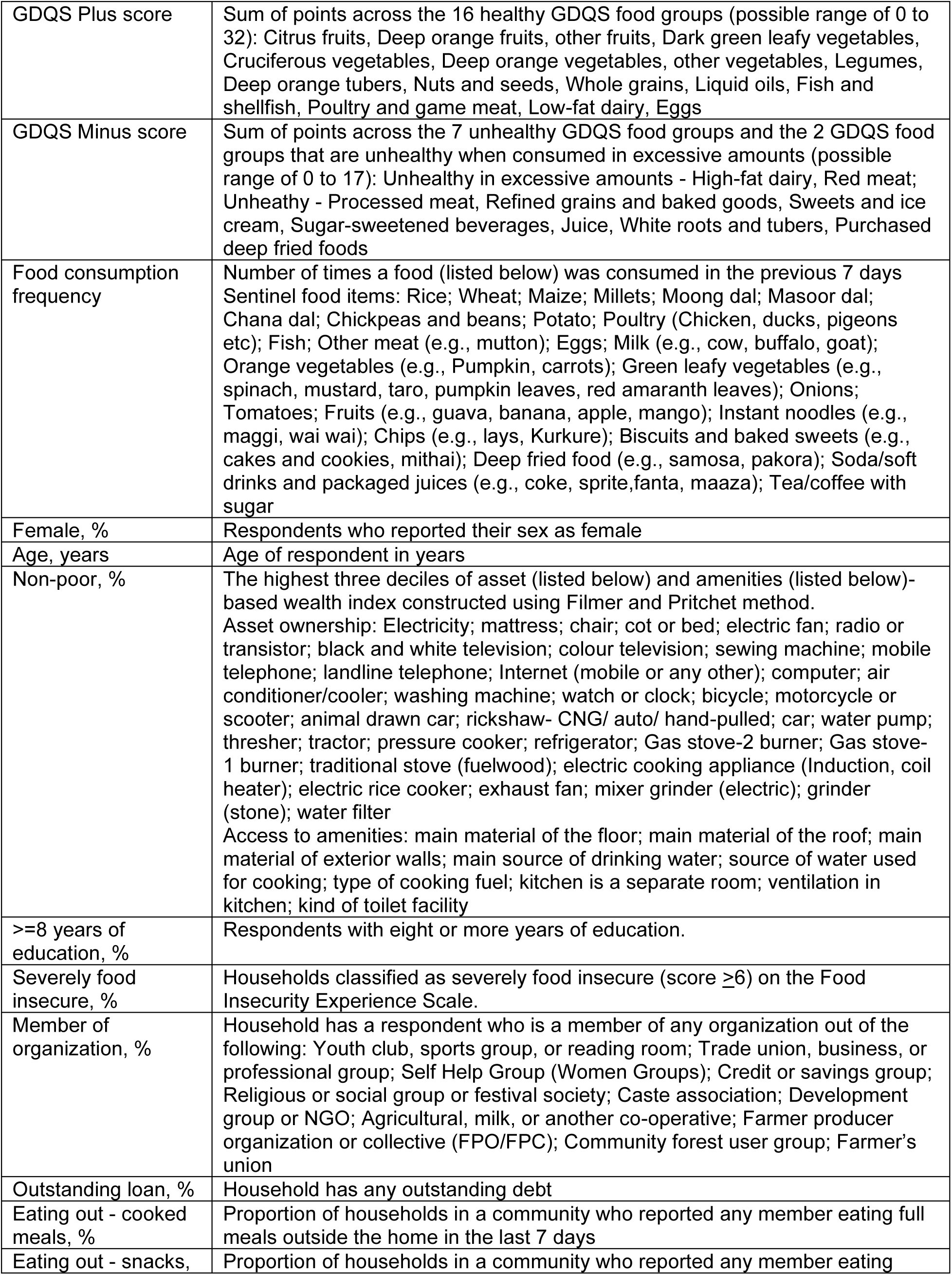
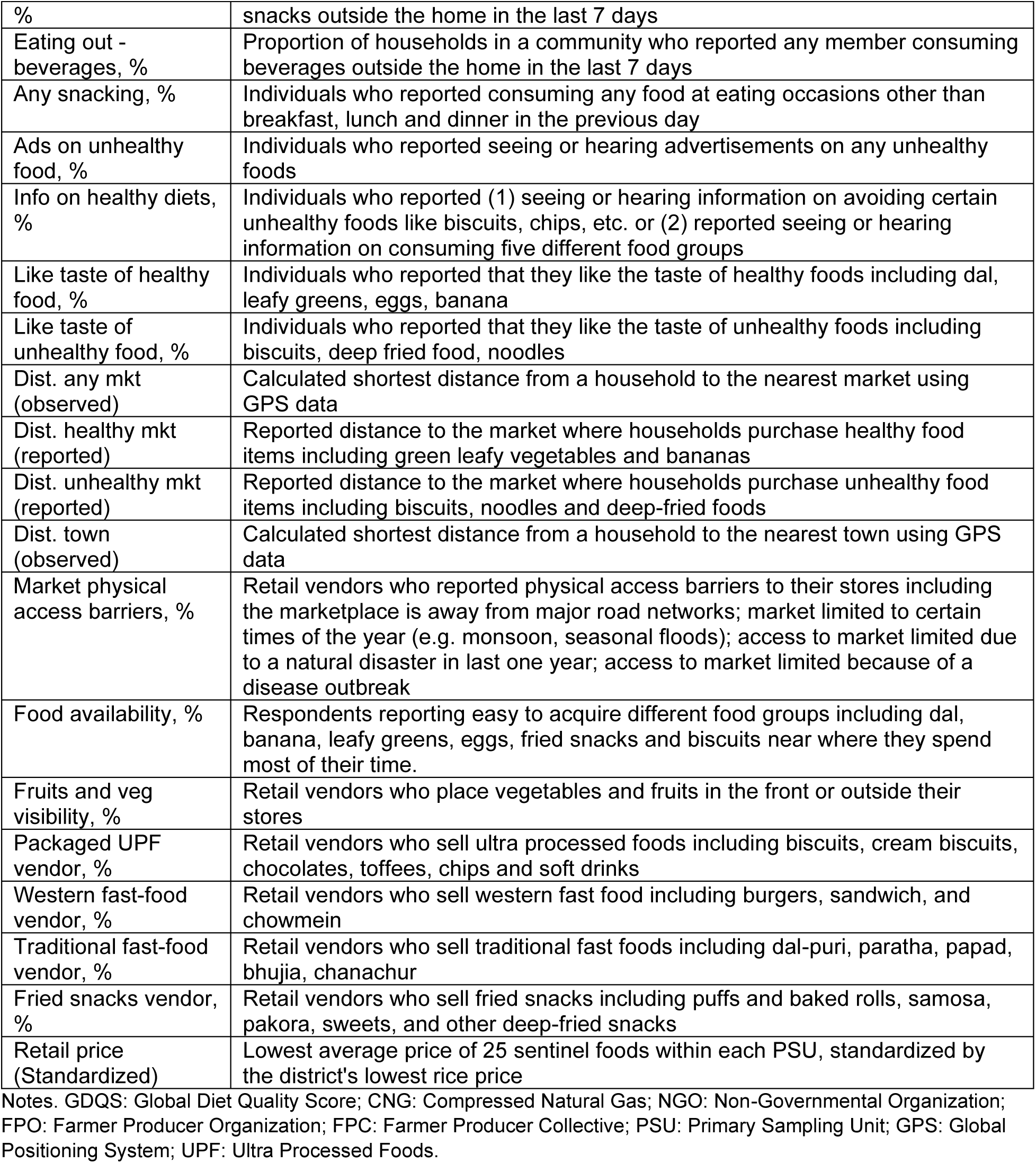
Definitions of individual, household and market level indicators used in the study.

### Food intake and diet quality

The Global Diet Quality Score (GDQS) app ^14^ was used to assess intake of GDQS food groups over the previous 24 hours. Respondents reported all foods and beverages consumed at home and outside, from waking until bedtime, across seven pre-defined eating occasions: pre-breakfast, breakfast, mid-morning, lunch, afternoon, dinner, and post-dinner. The GDQS-plus score was calculated by summing points from 16 healthy food groups (range: 0 to 32), while the GDQS-minus score summed points from 7 unhealthy food groups and 2 groups that become unhealthy in excess (range: 0 to 17). These two GDQS metrics were the primary dietary outcomes. See supplementary methods for details (S2). Secondary outcomes included the number of times 25 sentinel foods were consumed in the previous week from food frequency questionnaires (FFQ).

### Individual and household covariates

Individual-level covariates include respondent age (in years), sex (male or female), and years of education, which were included to capture demographic differences in dietary needs, preferences, mobility associated with aging, and nutritional knowledge related to healthier food choices. Tastes were captured by individual responses to questions regarding liking the taste of healthy (dal, eggs, bananas, and green leafy vegetables) and unhealthy (biscuits and fried snacks) foods. Snacking was measured using the GDQS application as a respondent eating at any occasion outside of breakfast, lunch, or dinner in the previous 24 hours.

Household-level covariates encompass household wealth deciles, severe food insecurity, and indebtedness to account for variations in the ability to afford a diverse and nutritious diet. Wealth deciles were constructed using Principal Component Analysis (PCA), following the methodology described by Filmer and Pritchett (2001). Severe food insecurity was calculated using the Food Insecurity Experience Scale (FIES) ^15^, which captures the extent of food-related constraints experienced by households.

Membership in organizations was also included as a covariate to account for the potential influence of community groups on food preferences and consumption behaviors. Detailed definitions for these indicators are available in **Table 1**.

### Food environment indicators

Food environment indicators calculated from the household survey include the availability of healthy and unhealthy foods, sources of exposure to advertisements promoting unhealthy foods, and sources of information on avoiding unhealthy foods, as reported by respondents. Availability indicators were scaled between 0 and 1, where 1 represents the availability of all food groups considered, and 0 indicates no food availability. The proportion of respondents who reported consuming full meals, snacks, or beverages outside the home was used to estimate eating out at the community level. Retail prices obtained from local vendors for all foods were standardized by using the lowest price of rice in a district as a numeraire or base value ^16^. Rice is a useful benchmark as it is the most common source of affordable calories in the region.

Food environment indicators calculated from the market survey include the visibility of fruits and vegetables in retail stores, the prevalence of vendors selling packaged ultra-processed foods, Western fast food, traditional fast food, and fried snacks. Additionally, community food prices obtained from retailers were used to capture affordability. To assess physical access to markets, several indicators were constructed to account for barriers and distances. First, market access barriers were proxied by vendors who reported obstacles to store access, such as marketplaces being far from major road networks, markets being open only during certain times of the year, and access issues due to natural disasters or disease outbreaks. Second, the shortest distance to markets and towns was calculated using the GPS coordinates of households and communities. Third, distances to markets selling healthy and unhealthy foods, as reported by respondents, were also included.

### Descriptive analysis

Several descriptive analyses were conducted. First, sample characteristics were described using means, proportions, and 95% confidence intervals for all variables within each district. Second, outcome distributions were visualized using kernel density curves by district. Third, the frequency and sources of advertisements and information on unhealthy foods were graphed across districts and household wealth categories. Fourth, district-level maps were created to assess geographic differences in the distribution and prevalence of unhealthy food vendors. Fifth, the distribution of eating out, snacking and wealth scores were visualized using kernel density curves to gauge normality and variation.

### Regression analysis

Two regression analyses were conducted. First, to analyze the effect of four key food environment domains—affordability, accessibility, availability, and desirability—on weekly food consumption, multivariate regressions were estimated using Equation (1). For individual *i*, in household *h*, community *c*, district *d*, and food *f*

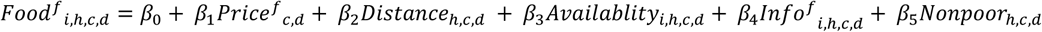

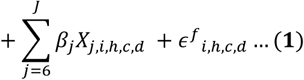

*Food* represents the weekly intake of 25 foods, *Price* (affordability) represents the community price of each food item, *Distance* (accessibility) represents the shortest distance to markets for each household measured in 10 km units, *Availablity* captures the reported level of healthy and unhealthy food availability in nearby markets, *info* (desirability) represents whether an individual heard messages promoting healthy food consumption (used as exposure for healthy foods) or saw ads promoting unhealthy food consumption (used as exposure for unhealthy foods) and *Nonpoot* (desirability and affordability) is a binary indicator for households in the top three wealth deciles, reflecting higher socioeconomic status. To interpret distance as accessibility we multiply the variable by −1 so it can be interpreted as proximity to markets. This transformation does not change the magnitude or significance of the coefficient, only the sign. The wealth variable serves as a proxy for desirability and affordability, capturing aspirational consumption patterns after adjusting for price effects because wealthier households are likely to consume more desired or aspirational foods due to greater purchasing power. *X*_*j,i,h,c,d*_ includes age, sex and district fixed effects. Standard error estimates are clustered at the household level to relax the independent errors assumption. All districts are assigned equal frequency weights to account for larger samples in Bangladesh relative to India and Nepal.

Second, to examine the association between multiple food environmental characteristics and diet quality, multivariate regressions were run using Equation (2). For individual *i*, in household *h*, community *c*, and district *d*,

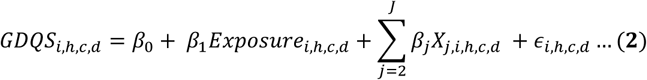

*GDQS* represents two diet quality metrics (GDQS-plus and GDQS-minus), *Exposure* refers to a specific food environment variable of interest, and *X*_*j,i,h,c,d*_ includes controls such as age, sex, education, wealth, severe food insecurity, organizational membership, outstanding loans, and district fixed effects. Standard errors are clustered at the household level. Regressions were conducted separately for each food environment variable to avoid biases due to multicollinearity.

## Results

### Individual diets and household demographics

**Table 2** and **Figure S3** show that GDQS-plus scores across districts ranged from 6.6 (SD 3.1) in Rangpur to 7.7 (SD 2.8) in Surkhet, indicating a low intake of healthy foods. GDQS-minus scores varied from 9.3 (SD 1.6) in Rajshahi to 11.7 (SD 1.7) in Nalanda, reflecting a moderate intake of unhealthy foods. Across all districts, 59-61% of respondents were female, with an average age of 29-32 years. Rajshahi and Banke had a higher proportion of non-poor households in terms of assets and amenities, and over one-third of respondents had 8 or more years of education across districts (**Table 2**). Severe food insecurity was lowest in Banke (2%) and highest in Rangpur (12%). In Nalanda, Banke, and Surkhet, more than 75% of households were members of an organization. Indebtedness was high across districts, with over 50% of households having an outstanding loan.

**Table 2.**
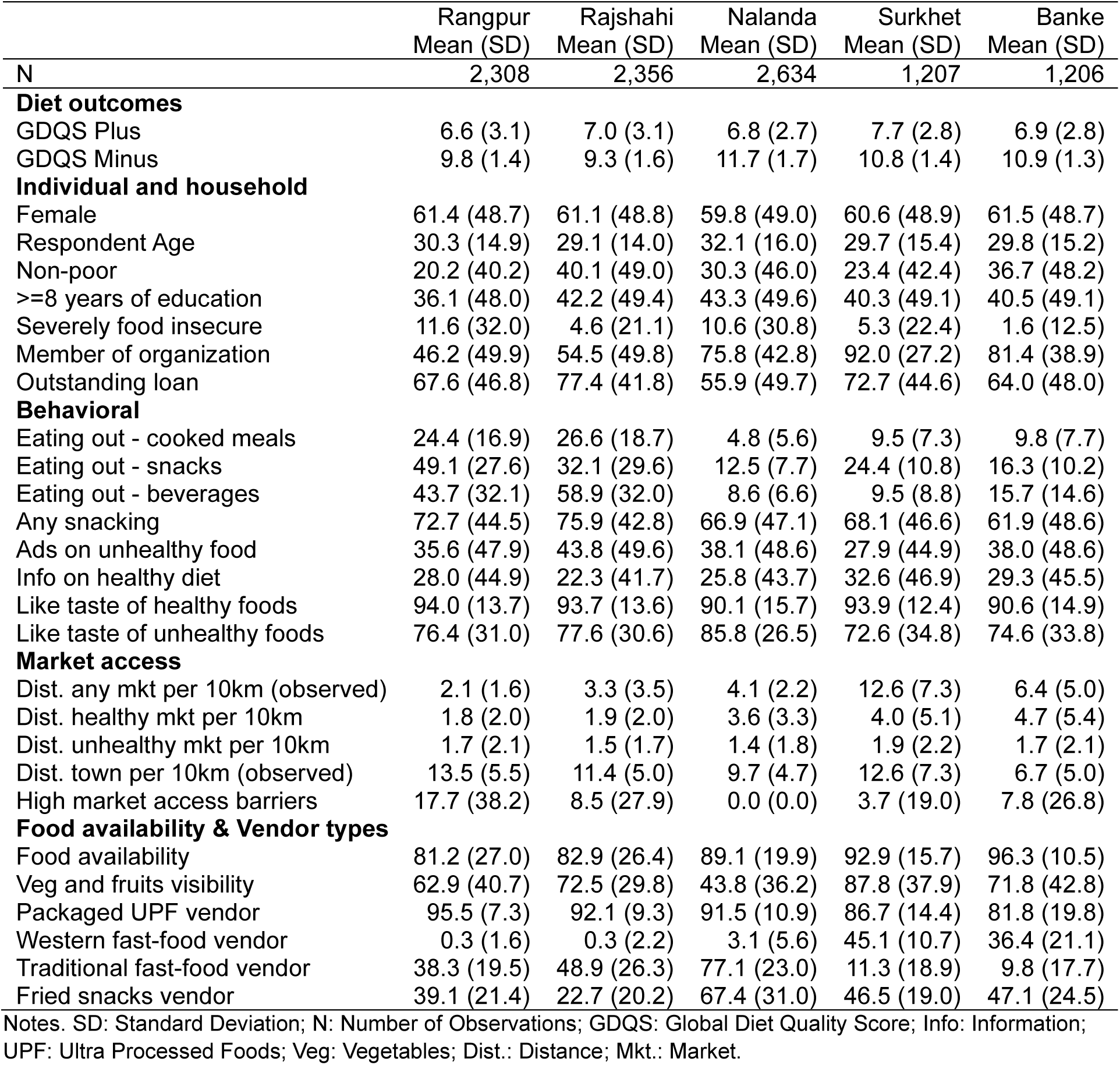
Sample characteristics for individual, household and market level indicators.

### Eating behaviors and exposure to advertising

Eating out—cooked meals (27%), snacks (32%), and beverages (59%)—was most prevalent in Rajshahi and Rangpur (**Table 2 and Figure S4**). Over one-third of respondents reported exposure to advertisements promoting unhealthy foods, while less than 20% reported seeing messages about avoiding unhealthy foods across districts. Most respondents indicated that advertisements for sugar-sweetened beverages were primarily encountered through media sources such as television and mobile phones (**Figure S5**). Additionally, respondents from non-poor households were more likely to be exposed to such ads (**Figure S5**). Snacking prevalence was notably high, with more than two-thirds of respondents reporting consumption of at least one meal outside of breakfast, lunch, and dinner (**Table 2 and Figure S4**).

### Local food environments

Reported food availability was high across districts (>80%) (**Table 2**). The visibility of fruits and vegetables relative to UPFs at retail outlets was lowest in Nalanda (44%) and highest in Banke (96%). Packaged UPF sellers were the most common type of retail vendors (>82%) (**Figure 1 and Table 2**). In Rangpur (38%), Rajshahi (49%) and Nalanda (77%), traditional fast-food sellers were prevalent, whereas western fast-food sellers were present in Surkhet (45%) and Banke (36%). Vendors selling fried snacks were common in all districts but highly prevalent in Nalanda (67%).

**Figure 1.**
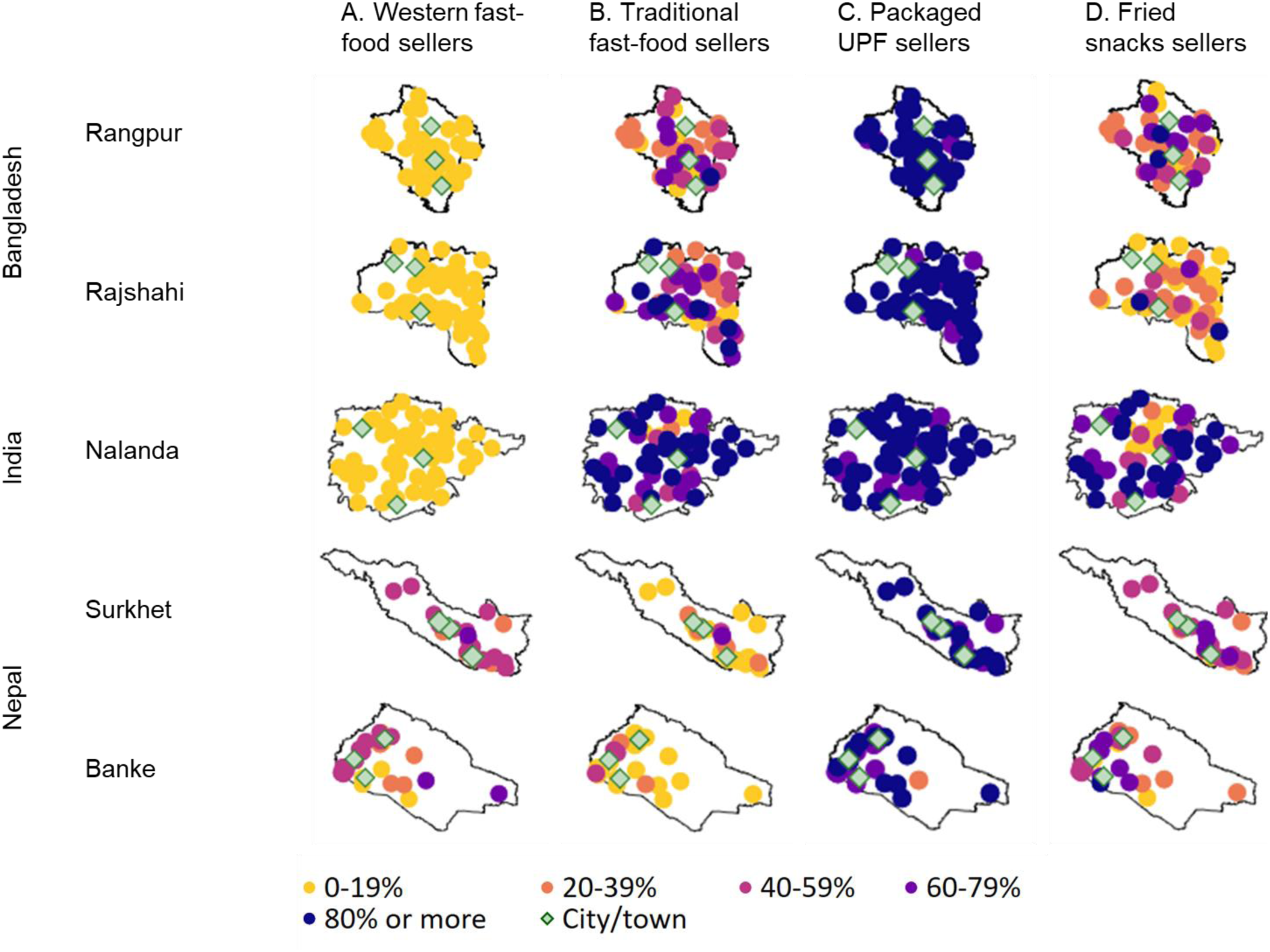
Vendor typology in communities across districts. Note. The figure presents spatial distribution of vendor typologies across five rural districts in South Asia: Rangpur, Rajshahi, Nalanda, Surkhet, and Banke. Each point on the map represent a village from study area.The variation in shading intensity represents the relative prevalence of each indicator, with the indicators divided into five quintiles and represented by the corresponding colors. The diamonds represent major cities or towns in each district. The vendor typologies depicted in the figure include: A. Western fast-food sellers, B. Traditional fast-food sellers, C. Packaged UPF sellers and D. Fried snacks sellers. The definitions of these indicators are available in **Table 1**. UPF: Ultra-processed food.

In Rangpur, Rajshahi, and Nalanda, the measured distance to the nearest market ranged from 2 to 4 km, closely aligning with the distances reported by households (**Table 2**). However, in Banke and Surkhet, the measured distance to markets was significantly higher (6-13 km) compared to reported distances (2-5 km). Across all districts, the distance to unhealthy food vendors was consistently shorter than the distance to healthy food vendors. The nearest town was, on average, 7 to 14 km away. Market access barriers were highest in Rangpur (18%) and Rajshahi (9%), but relatively low in the other districts.

### Affordability, accessibility, availability, desirability, and food choice

**Table 3** presents the regression estimates derived from Equation (1). Living close to markets, as a measure of accessibility, was associated with higher weekly consumption of fresh fruits and vegetables but lower consumption of fish and staples like wheat and maize (p<0.001). Increased reported food availability in markets consistently predicted higher weekly consumption of unhealthy foods such as chips, baked snacks, fried foods, sugary beverages, and tea (p<0.001), indicating that these rural markets resemble “food swamps.” Higher retail food prices (adjusted for wealth) were negatively associated with most food groups, with significant reductions observed for rice, wheat, poultry, meat, tomatoes, and sugary beverages (p<0.001). Messaging designed to increase desirability of healthy (through information) and unhealthy (through ads) foods showed large, positive associations with consumption of wheat, fish, eggs, vegetables, fruits, baked goods and tea (p<0.001). Desirability (revealed preference) and affordability (higher wealth), proxied by non-poor households (adjusted for prices), were strong predictors of higher consumption of nearly all foods (p<0.001), except for maize, millets, and potatoes.

**Table 3.** Association between indicators of affordability, accessibility, availability and desirability with weekly frequency of consuming various foods.

|  | Accessibility | Availability | Affordability |  | Desirability |
| --- | --- | --- | --- | --- | --- |
|  | Proximity to market, per 10 km | High food availability, lowest versus highest | Retail prices standardized by lowest rice price, rupees/taka/kg | Non-poor, highest wealth tertile | Seen or heard messages about healthy/unhealthy foods |
| Rice | 1.01*** | 0.87*** | -0.32*** | -0.92*** | -0.21 |
| Wheat | -1.19*** | 0.04 | -0.37** | 0.98*** | 0.30* |
| Maize | -0.47*** | 0.07 | 0.26* | -0.12* | -0.08 |
| Millets | -0.04 | 0.06 | 0 | -0.03 | 0.08 |
| Potato | 0.70* | 0.47 | 0.13 | -0.02 | -0.23 |
| Moong dal | -0.02 | 0.12 | -0.13 | 0.27*** | 0.16** |
| Masoor dal | 0.70** | -0.59* | 0.54*** | 0.73*** | 0.13 |
| Chana dal | -0.16 | -0.01 | 0.31** | 0.40*** | 0.15* |
| Chickpeas | 0.70*** | 0.58*** | -0.05 | 0.35*** | 0.02 |
| Poultry | 0.2 | 0.14 | -0.01** | 0.45*** | 0.07 |
| Fish | -0.82*** | -0.68* | 0.01 | 0.70*** | 0.38*** |
| Other meat | 0.05 | 0.11 | -0.08*** | 0.36*** | 0.05 |
| Eggs | 0.24 | 0.40** | -0.2 | 0.65*** | 0.31*** |
| Milk | -0.48* | -0.08 | -0.1 | 1.08*** | 0.22* |
| Orange veg | 0.03 | 0.1 | -0.02 | 0.59*** | 0.13** |
| Leafy veg | 0.50** | -0.19 | 0.17 | 0.35*** | 0.32*** |
| Onion | 1.47*** | 0.57 | 1.02 | 1.28*** | 0.73*** |
| Tomato | 1.65*** | -0.08 | -1.86*** | 1.70*** | 0.73*** |
| Fruits | 0.41** | 0.56** | -0.10* | 1.00*** | 0.24** |
| Noodles | -0.23** | 0.11 | 0 | 0.19*** | 0.17*** |
| Chips | 0.05 | 0.26*** | -0.01 | 0.14*** | 0.04 |
| Baked | 0.04 | 0.89*** | 0 | 0.50*** | 0.16* |
| Deep fried | 0.08 | 0.69*** | -0.11** | 0.34*** | 0.13** |
| Sugary beverage | 0.22** | 0.22*** | 0.03** | 0.36*** | 0.13*** |
| Tea/coffee with sugar | 0.31 | 2.32*** | -1.75*** | 0.96*** | 0.22 |

Notes: \* $p < 0.1$ , \*\* $p < 0.05$ , \*\*\* $p < 0.01$ . Outcomes are measured as the number of times a food was consumed in the previous week. Coefficients from multivariate regression (equation 1) show associations between weekly food consumption frequency and food environment indicators. Accessibility measures the shortest distance from household to nearest market using GPS coordinates (per 10km). Availability represents the proportion of households where respondents reported easy to acquire food groups (dal, banana, leafy greens, eggs, fried snacks and biscuits) near where they spend most of their time. Price variables use the lowest average price of listed sentinel foods within each village, standardized by the district's lowest rice price. Non-poor represents the highest wealth tertile households. For rice to fruits, 'seen or heard messages' refers to health messaging about avoiding processed foods or consuming five food groups; for noodles to tea/coffee, it refers to messages about unhealthy foods and packaged drinks. Blue (red) cells indicate positive (negative) associations, with color intensity proportional to coefficient magnitude. Explanatory variables included in the regression are age, sex, and district fixed effects. Standard error estimates are clustered at the household level.

### Food environment, socioeconomic, and behavioral drivers of diet quality

**Figure 2** presents the regression estimates derived from Equation (2). Snacking emerged as the strongest predictor of diet quality, showing a positive association with both healthy food intake (+1.15 GDQS-plus) and unhealthy food intake (−0.67 GDQS-minus). Liking the taste of healthy and unhealthy foods was also strongly associated with an increase in healthy (+0.98 GDQS-plus) and unhealthy (−0.32 GDQS-minus) food consumption. Consuming cooked meals outside the home was linked to a higher intake of unhealthy foods (−0.28 GDQS-minus), while drinking beverages outside the home predicted lower consumption of healthy foods (−0.47 GDQS-plus). Exposure to food advertising was associated with an increase in both unhealthy food consumption (−0.09 GDQS-minus) and healthy food consumption (+0.38 GDQS-plus).

**Figure 2.**
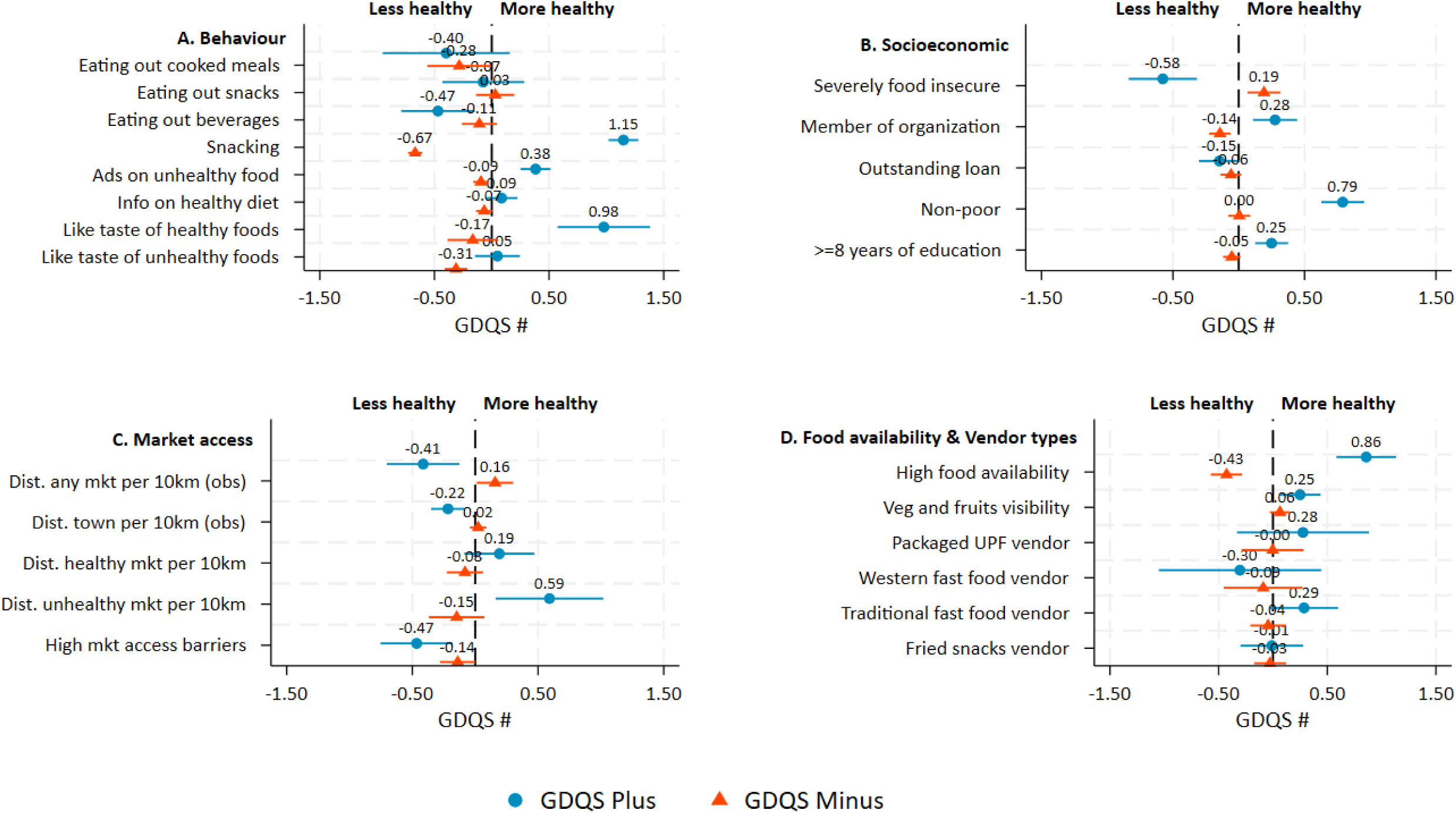
Food environment, socioeconomic, and behavioral drivers of diet quality. Notes: The figure presents coefficients with 95% confidence intervals from multivariate regression models (see equation 2) examining association between diet quality indicators, GDQS plus (blue circles) and GDQS minus (orange triangles) across four domains: A. Behaviour, B. Socioeconomic, C. Market access, D. Food availability & Vendor types. Scores with positive values indicate healthier associations and negative values indicate less healthy associations. Zero represents no association. Models A, C and D are adjusted for all predictors from model B. i.e. age, sex, household food insecurity, member of organization, outstanding loan, wealth, education, and district fixed effects. Standard error estimates are clustered at the household level. GDQS: Global Diet Quality Score; UPF: Ultra Processed Foods; Dist.: Distance; obs: observed measurements per 10km radius; mkt: Market.

Among socioeconomic covariates, being a member of an organization (+0.28), being non-poor (+0.79), and having 8 or more years of education (+0.25), was significantly associated with higher intake of healthy foods (GDQS-plus). Expectedly, severe food insecurity predicted both lower healthy (−0.58 GDQS-plus) and unhealthy (+0.19 GDQS-minus) food intake, indicating a severe lack of resources.

Measured distance to markets (per 10 km) was associated with lower consumption of both healthy foods (−0.41 GDQS-plus) and unhealthy foods (0.16 GDQS-minus). In contrast, a greater reported distance to markets selling unhealthy foods was linked to significantly higher healthy food consumption (+0.59 GDQS-plus). High barriers to market access were associated with significantly lower GDQS-plus and GDQS-minus scores, indicating reduced overall food intake. Greater food availability predicted both higher consumption of healthy foods (+0.86 GDQS-plus) and unhealthy foods (−0.43 GDQS-minus). Additionally, higher visibility of fruits and vegetables in retail outlets was linked to increased healthy food consumption (+0.25 GDQS-plus), as was the presence of traditional fast-food vendors (+0.29 GDQS-plus).

## Discussion

The examination of rural food environments across South Asian districts has four key findings with significant implications for public health nutrition and policy. First, suboptimal diets in these regions are characterized by low consumption of healthy foods and moderate consumption of unhealthy foods, reflecting an urgent need to promote more balanced dietary patterns. Second, affordability emerges as the primary driver of healthy food intake, while the interplay of affordability, desirability, and availability drives unhealthy food consumption, underscoring the complex socio-economic determinants of dietary behavior. Third, rural food environments are undergoing rapid urbanization, as evidenced by eating out, exposure to targeted food messaging, shifts in tastes and preferences, accessibility to markets, and the presence of diverse fast-food options, all of which significantly influence diet quality. Finally, snacking between meals stands out as the most significant predictor of both healthy and unhealthy food consumption, highlighting its dual role in shaping dietary outcomes. This study highlights the urgency of developing comprehensive policies that align economic, behavioral, and environmental factors to promote healthier rural food environments.

Strengths of the study include the examination of multiple dimensions of rural food environments: affordability, accessibility, availability, and desirability, among others. The study uses robust analytical techniques, controlling for key covariates, including socioeconomic, demographic, and geographic factors, to reduce threats from omitted variables. The study leverages primary data to measure the unique characteristics of South Asian food environments, making the findings highly relevant to policymakers and researchers in similar settings. The inclusion of diverse rural districts across South Asia enhances the generalizability of the findings to similar LMIC settings. However, our study is not without limitations. The cross-sectional nature of the analysis makes it difficult to infer causal relationships due to threats from residual confounding. Thus, the estimates from the regression models should be interpreted as associations rather than causal. In addition, as self-reported recall based dietary data is subject to inaccuracies, participants’ 24 hour or 7-day dietary recall may underestimate or overestimate consumption, especially for less frequent food items. Further, the study provides a static view of food environments and dietary behaviors, missing seasonal variations in food availability or affordability. Nevertheless, these results provide a robust foundation for future research, particularly studies that explore the direct linkages between market data and household consumption patterns to further enrich our understanding of rural food environments.

One critical feature of unhealthy diets in our analysis was the low consumption of healthy foods, often overshadowed by the widespread availability and affordability of ultra-processed, nutrient-poor options. Affordability remains a key barrier in South Asia, where nutrient-dense foods such as fruits, vegetables, and protein rich foods are often priced out of reach for many households ^17,18^. Addressing affordability challenges requires innovative policy solutions such as integrating nutrient-rich foods into school feeding programs, expanding safety nets like food or cash transfers, or piloting food voucher programs specifically for enabling consumers to access fruits and vegetables while stimulating local markets ^19,20^. In addition, countries in South Asia struggle with horticultural productivity and supply chain issues for fruit and vegetables; warranting urgent agricultural investments ^21^. Further, the promotion of healthy food desirability is not feasible without widespread awareness across schools, consumer associations, producer co-operatives, women collectives and health centers. Evidence on the choice and efficacy of these platforms can provide valuable insights for influencing food environments in South Asia.

Affordability, desirability and availability were identified as key drivers of unhealthy food consumption, as attractive packaging, taste appeal, and widespread accessibility make UPFs and SSBs highly appealing ^22^. The abundance of these products, often positioned prominently in high-traffic areas, further normalizes their consumption ^23^. Interventions to address this include regulating product placement in retail stores to limit visibility and introducing warning labels to highlight the health risks of UPFs ^24^. Agreeing with prior research, our analysis shows that visibility of fresh produce at locations around stores is associated with higher GDQS+ scores. To curb unhealthy consumption, several countries have strengthened the regulatory framework for food packaging, labelling, and marketing to influence perception and desirability of unhealthy food products. Enforcing front-of-pack nutrition labelling (FOPNL) is widely practiced for informing consumers regarding nutritional quality of packaged foods and nudging them away from unhealthy products. Implementing FOPNL throughout South Asia is a clear ask. Engaging with alternative FOPNL approaches (nutri-score, health star rating, traffic lights, warning labels etc.) is a priority research area to establish their relative effectiveness. Curbing unhealthy consumption also requires focus on fiscal instruments such as taxes on SSBs and/or other unhealthy foods to reduce their affordability and incentivize healthier choices ^25^. Application of tax slabs on various types of food high in fat, sugar and salt (HFSS) is evident across various contexts even though there is limited evidence on tax incidence and market distortions associated with such measures.

Our study suggests that snacking can have large positive and negative impacts on diets in low-income settings. Snacking can serve as an entry point to encourage dietary diversity, particularly in low-income settings where main meals often consist of repetitive food choices in a single day. Snacks provide an opportunity to introduce nutrient-rich foods like fruits, vegetables, and nuts, which may not feature in regular meals ^26^. Interventions can promote diverse snacks through social marketing or other approaches; this is a key area for potential future research and might offer more feasibility than changing entire meal patterns. For example, community and school level education campaigns focusing on dietary diversity and simple snack recipes can encourage variety. Although the World Health Organization (WHO) has developed specific recommendations to reduce marketing exposure of children and adolescents toward unhealthy food and beverages, South Asian countries still have huge exposure to advertisements through television as well as print and digital platforms. High volume of advertisements is predominantly observed for HFSS foods whereas limited promotion of healthy food options is noted ^27,28^. Research on food marketing can provide insights into advertisement regulations on unhealthy food and beverages targeting children and adolescents.

### Conclusion

South Asia’s rural food environments increasingly resemble “food-swamps” where traditional foods coexist with ultra-processed nutrient-poor options^23^. The interplay of affordability, availability, and desirability shapes diets both in enriching and detrimental^29^. Our findings highlight the need for tailored interventions: addressing the affordability of nutrient-dense foods, enhancing the visibility and accessibility of fresh produce, and reimagining food marketing and financial incentives or disincentives to empower healthier choices. Snacking offers a potential entry point to foster dietary diversity while curbing the normalization of unhealthy consumption. These insights call for a multisectoral approach, uniting policymakers, producers, and consumers. The future of South Asia’s food systems lies in ensuring that modernization nurtures rather than erodes the foundations of rural diets.

## Data Availability

The datasets analyzed during this study are publicly available and can be accessed at https://www.cgiar.org/news-events/news/open-access-agrifood-system-data-from-4000-households-across-bangladesh-india-and-nepal/ and https://dataverse.harvard.edu/dataverse/IFPRI/?q=%22TAFSSA+District+Agrifood+Systems+Assessment%22+AND+%22Market%22+AND+%22Retail%22

## Supplementary tables and figures

### S1. Household survey data collection and sampling

Survey locations were selected based on being hot spots for challenges related to poverty, climate, malnutrition and social inclusion, and having CGIAR staff presence. The surveys are representative at the district level. The sampling approach and sample size determination followed the example of India’s equivalent of the Demographic and Health Surveys, the National Family Household Survey (NFHS), a population-based survey representative at the district level most recently conducted in 2019-2021. We erred on the higher end of the number of households per district in NFHS and decided on a sample size of 1000 households per district for our survey. In Nepal, relative to Bangladesh and India, districts were much less populated, thus we included 500 households per district in Nepal. The primary sampling unit (PSU) was villages (in Bangladesh and India) and wards (in Nepal). PSUs were selected from National Census datasets from each country, using a Probability Proportional to Size (PPS) sampling strategy. In the first stage of sampling, villages or wards were selected at random, with a probability proportional to the number of households that reside in the PSU. In the second stage, an equal number of households were selected randomly from each PSU, roughly 20 households per PSU. The sampling frame included all rural households that included adolescent members. The rationale for only including households with adolescents was to better understand adolescents’ consumption behavior and their roles in agrifood systems, which will be presented in a forthcoming paper.

The adult female respondent was identified as the female household member primarily responsible for managing the household. The adult male respondent was identified as the male household member primarily responsible for agricultural activities. Three survey firms supported data collection: Data Analysis and Technical Assistance in Bangladesh, Kabil Professional Services in India, and Institute of Integrated studies in Nepal. After explaining the survey’s objectives and procedures, enumerators obtained verbal informed consent from respondents. Participation in the survey was voluntary, and participants were free to end interviews at any time by informing survey staff.

### S2. GDQS data collection procedure

The Global Diet Quality Score (GDQS) app (Moursi et al. 2021) was used to capture intake of GDQS food groups in the previous 24 hours. The GDQS app was pilot tested and the GDQS food database was adapted to each survey context. The list of over 6000 food items and the app instructions and interface were translated to Bangla, Hindi, and Nepali. Enumerator training was conducted separately in each country using the GDQS app and plastic cubes (described more below). The GDQS application was then pre-tested and any further context-specific adjustments were incorporated. The GDQS interview start time and end time were recorded as the date on which the interview took place followed by the time in hours, minutes, and seconds. The GDQS interview duration was calculated in minutes as the difference between the end time and the start time. The mean duration of dietary recall using the GDQS app was 32 minutes per respondent (a mean of 35 minutes in Banke, 27 minutes in Surkhet, 50 minutes in Nalanda, 24 minutes in Rajshahi, and 23 minutes in Rangpur).

Seven steps were used to collect data on tablets using the GDQS app.

- Step 1 involved collecting demographic information about the respondent.
- In step 2, respondents were asked about everything they ate or drank at home and outside the home on the previous day, from the time they woke up until the time they went to bed and didn’t eat or drink anything more. This information was recorded as foods/dishes/beverages consumed during the following pre-determined list of seven eating occasions: pre-breakfast, breakfast, mid-morning, lunch, afternoon, dinner, and post-dinner.
- For each eating occasion, all foods and drinks reported by the respondents were recorded. In case a respondent consumed a mixed dish (a dish prepared using multiple ingredients), they were requested to list the main ingredients of each mixed dish in step 3. Information about spices used in mixed dishes was not collected because spices do not get categorized as a GDQS food group, and the quantity of spices used is often unknown and difficult to estimate.
- In step 4, additional information on certain foods was collected to help classify these ingredients into the GDQS food groups. For instance, if a respondent mentioned that they ate bread, there was a follow-up to question asking whether it was white or dark brown to classify bread either in the refined or the whole grains GDQS food group.
- Step 5 checked whether any of the reported foods were purchased deep fried.
- Step 6 recorded any sugar or caloric sweeteners consumed that might have been missed in the open recall in step 2.
- In step 7, respondents were requested to visualize the total amount of foods and beverages consumed belonging to the same food group. A set of ten 3D-printed numbered plastic cubes in a range of predetermined sizes was provided to aid the visualization process. For each food group, the enumerator read back to the respondent all foods belonging to the same food group as classified by the application (based on the open recall in step 2) and asked them to point to the cube size that came closest to the amount of all foods combined belonging to the same food group. For example, if they consumed an apple and a banana (both in the ‘other fruit’ food group) yesterday, they were asked to select the cube number corresponding to the amount of apple and banana combined total volume of the apple plus the banana. Cube sizes were used as a measure of consumed quantity being below, equal to, or above food group–specific cutoffs established in grams, as described in Moursi et al. 2021.

**Figure S3.**
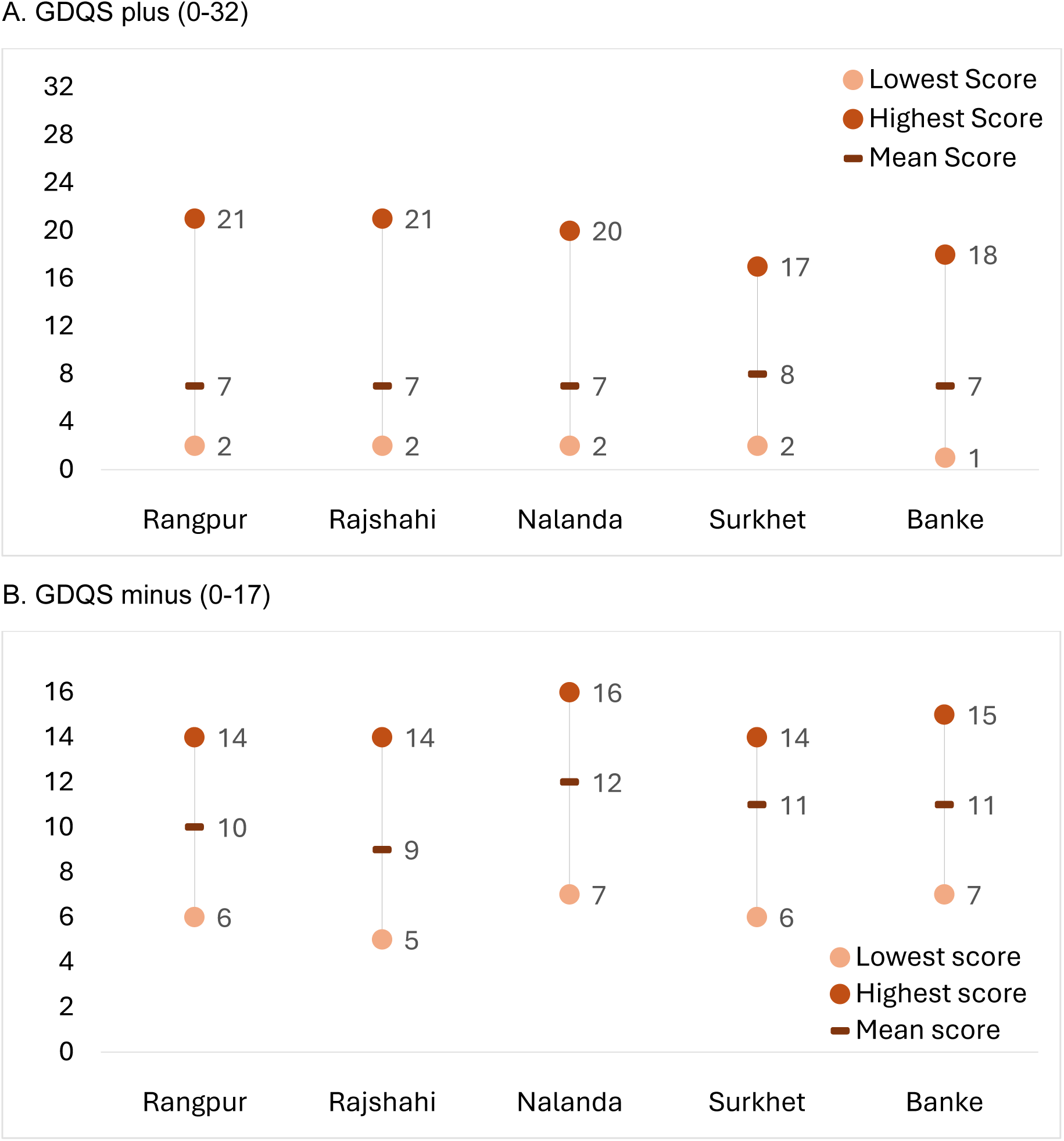
Diet quality scores across districts. Notes. The figure provides an overview of diet quality variations across five rural South Asian districts—Rangpur, Rajshahi, Nalanda, Surkhet, and Banke. Panel A represents GDQS plus scores (range: 0–32), indicating higher healthy food intake as the score increases. Panel B represents GDQS minus scores (range: 0–17), indicating lower unhealthy food intake as the score increases. For each district, the lighter point indicates the lowest score, the darker point indicates the highest score, and the horizontal bar represents the mean score. The y-axis denotes the respective score values.

**Figure S4.**
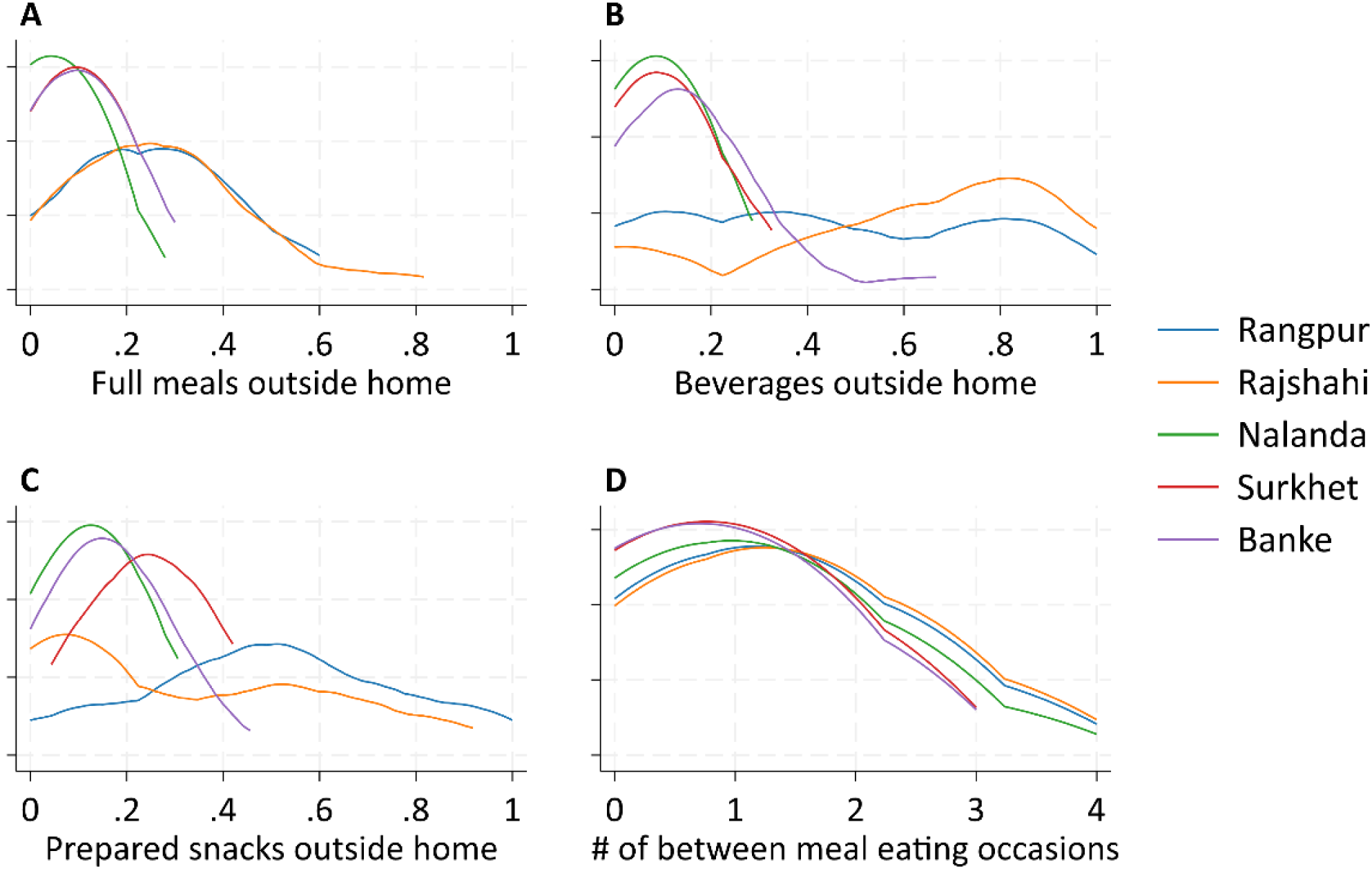
K-density curves showing the distribution of eating behavior by districts. Notes. The figure illustrates the distribution of food consumption behaviors across five rural South Asian districts— Rangpur, Rajshahi, Nalanda, Surkhet, and Banke. The x-axis in each panel represents the proportion (A-C) or count (D), while the y-axis represents the density or frequency of the food consumption behaviors across the study sample. The curves depict patterns for each district, distinguished by different colors as outlined in the legend.

**Figure S5.**
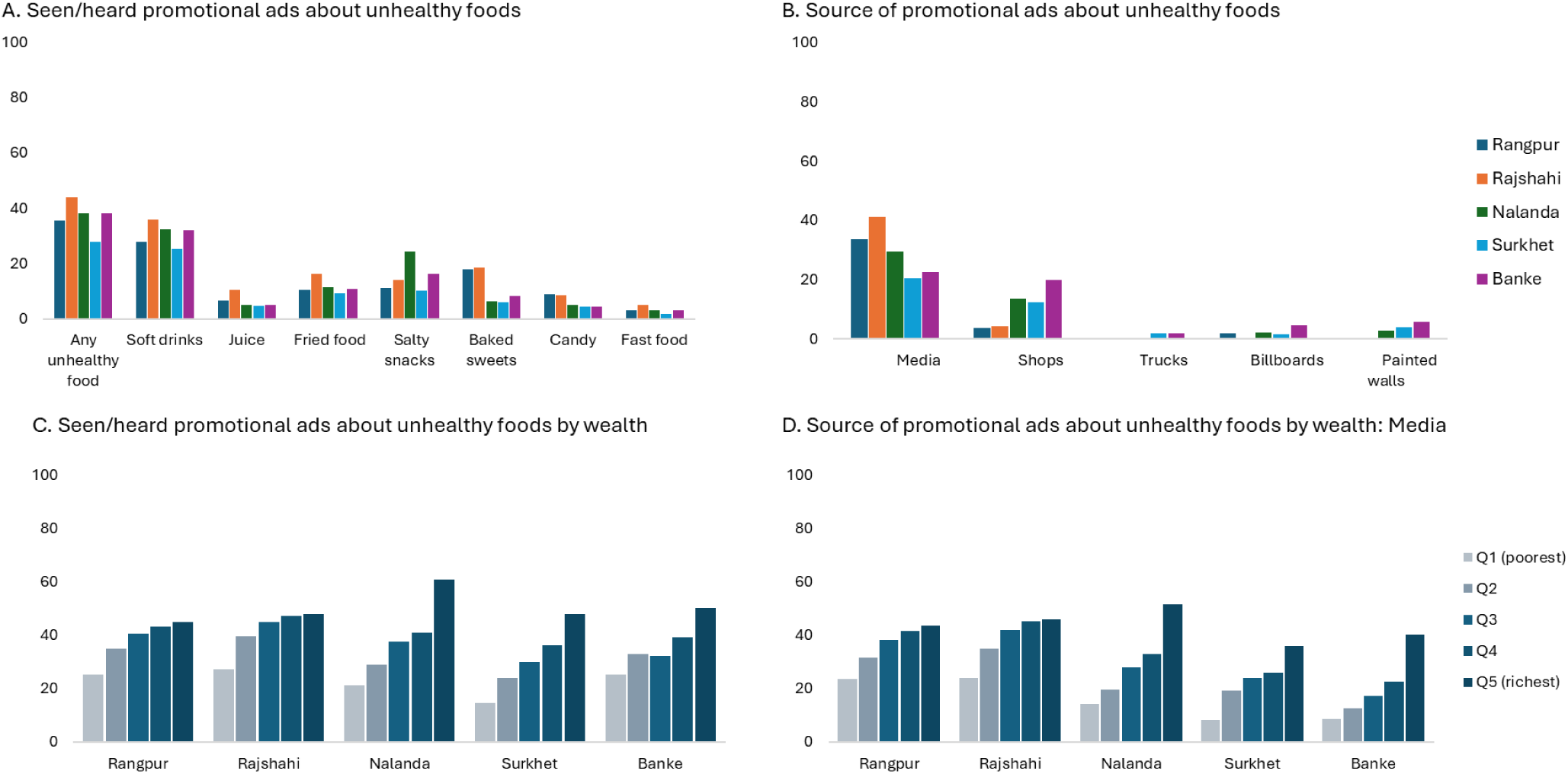
Exposure to promotional advertisements about unhealthy foods. Notes. The figure illustrates exposure to advertisements for unhealthy foods across five South Asian districts— Rangpur, Rajshahi, Nalanda, Surkhet, and Banke. Panel A presents respondents’ reported exposure to promotional ads for various unhealthy foods. Panel B highlights the sources of these advertisements, with respondents allowed to select multiple options from a provided list. Panel C shows the proportion of respondents exposed to unhealthy food ads based on household asset ownership (Q1: lowest wealth quintile, Q5: highest). Panel D illustrates the proportion of advertisements seen or heard through media, further categorized by household asset ownership across districts.

